# Online tests for sexually transmitted infections – Friend or Foe? An analysis of providers in the United Kingdom

**DOI:** 10.1101/2021.07.01.21259784

**Authors:** Eleanor Clarke, Paddy Horner, Peter Muir, Katy M. E. Turner, Emma M. Harding-Esch

## Abstract

**Objectives:** Online testing for sexually transmitted infections (STIs) may contribute to overcoming barriers to traditional testing such as stigma and inconvenience. However, regulation of these tests is lacking, and the quality of services is variable, with potential short- and long-term personal, clinical and public health implications. This study aimed to evaluate online tests available in the UK against national standards.

**Methods:** Providers of online STI tests (self-sampling and self-testing) in the UK were identified by an internet search of Google and Amazon (June 2020). Website information on tests and care was collected, and further information requested from providers via an online survey, sent twice (July 2020, April 2021). The information obtained was compared to British Association for Sexual Health and HIV (BASHH) guidelines for diagnostics and standards of STI management.

**Results:** 31 providers were identified: 13 self-test, 18-self-sample, and two laboratories that serviced multiple providers. Seven responded to the online survey. Many conflicts with national guidelines were identified, including: lack of health promotion information, lack of sexual history taking, use of tests licensed for professional use only marketed for self-testing, inappropriate infections tested for, incorrect specimen type used, and lack of advice for post-diagnosis management.

**Conclusions:** Very few online providers met the BASHH national STI management guidelines standards that were assessed, and there is concern that this will also be the case in areas that were not covered by this study. For-profit providers were the least compliant, with concerning implications for patient care and public health. Regulatory change is urgently needed to ensure that online providers are compliant with national guidelines to ensure high-quality patient care, and providers are held to account if non-compliant.

**Key message box:**

- Online providers help overcome many barriers to STI testing and are increasingly popular, but quality of services is not assured
- Many online testing services, particularly for-profit providers, did not comply with national guidelines
- Substandard services can lead to serious personal, clinical and public health
- implications, such as inappropriate testing, inappropriate antimicrobial prescribing, unnecessary emotional distress and missed diagnoses
- Regulatory change is required to ensure online providers comply with national guidelines and are held to account when they do not

## INTRODUCTION

Sexually transmitted infections (STIs) are an increasing public health problem in the world,[1] including the United Kingdom (UK).[2] Early diagnosis is a core intervention for guiding appropriate management, thus reducing the risk of antimicrobial resistance (AMR) emergence, preventing reproductive sexual health (RSH) sequelae, and reducing onward transmission.[1] Therefore, access to validated and approved testing services is vital. Tests for self-directed use available to purchase online (henceforth referred to as ‘online tests’) are increasingly popular, especially with the advent of the COVID-19 pandemic.[3] Even pre-COVID-19, online tests were widely viewed as an asset to public health, with studies demonstrating they can overcome barriers such as stigma and inconvenience,[2,4] and were the second most frequent testing service type in the UK’s National Chlamydia Screening Programme (NCSP) in 2019.[2] Online tests come in two main forms: self-sampling, where the user can order a kit, take a specimen independently and then post the sample to the laboratory with results received through a text, letter or online portal;[5] and self-testing, where the user collects a specimen, conducts the test themselves, and interprets the result in private.[5]

However, drawbacks to online testing services have also been widely reported. Barriers for use include technical ability, language, and health or digital literacy.[4] Lack of interaction with a health professional may also be a worry to users and can result in improper management of infections and follow-up care.[6] In the UK, standards for providers of sexual health services are published by the British Association for Sexual Health and HIV (BASHH) and Faculty of Sexual and Reproductive Healthcare (FSRH).[7,8] Within these standards is the stipulation that services must include health promotion and prevention interventions and correct signposting information,[7] however, these may not always be present,[9] leaving users vulnerable to making misinformed decisions. Additionally, private testing may result in data not being captured by national surveillance systems, posing issues for epidemiological monitoring.[6,10]

A key concern relating to online testing is the quality of the tests themselves. Although product regulatory standards such as CE-marking are often used to assess product quality,[11] this may not always indicate good performance.[12] Poor diagnostic accuracy can lead to false-positives resulting in unnecessary treatment, with resulting AMR risk and relationship implications, and false-negatives enabling further transmission and RSH sequalae.[13] Furthermore, some testing panels include infections that STI guidelines recommend should not be routinely tested for,[14,15] resulting in similar consequences to false-positive test results.

In this study, we assessed whether UK patients accessing online STI tests are receiving quality of care consistent with diagnostic standards. First, we identified and characterised online test providers in the UK, before comparing them to BASHH diagnostic guidelines [16] and BASHH standards for STI management.[7]

## METHODS

Ethical approval was granted by the MSc Research Ethics Committee of the London School of Hygiene & Tropical Medicine (reference number 22195).

### Internet search of providers

To identify tests available in the UK, a structured search of Google and Amazon was completed on 27^th^ June 2020. Full details of the search are available in Supplementary Table 1. To reduce unrelated results, a combination of smaller searches was used, and the results pooled. Searches aimed to replicate the consumer experience, so layman’s terms were used.

**Table 1:** A summary of recommendations from the British Association of Sexual Health and HIV (BASHH) standards for the management of sexually transmitted infections (STIs).[7] These were used to assess whether online providers were providing an adequate standard of care to patients

| Standard | BASHH Standards for STI Management |
| --- | --- |
| 1: Test audience and pre-test process | Online services are not appropriate for people with genital symptoms or symptoms of sexual infection<br>There should be the recording of a medical and sexual history, including use of contraception and drug and alcohol risk assessment, whether taken by a healthcare professional or self-completed. |
| 2: Test process | Specimens for STI testing including as a minimum those for chlamydia, gonorrhoea, syphilis and HIV from all relevant exposed sites.<br>Specimens for microbiological testing obtained during the examination should be in line with national guidance. This should include samples from extra-genital sites if indicated in the sexual history.<br>If home sampling or testing kits are provided, it is the providers' responsibility to ensure there are clear and comprehensive written instructions on how to use the kit and how to access follow up and support if necessary |
| 3: Health information and signposting | The STI tests offered to individuals should be explained clearly, including what they are for, how samples are taken, the limits of each test i.e. window periods<br>Health promotion and prevention interventions including encouragement of safer sex behaviour and condom usage.<br>Providers should ensure that appropriate advice is given about effective preventative interventions such as post exposure prophylaxis |
| 4: Follow Up/Treatment | Have clear agreed care pathways in place to ensure people have access to appropriate care based on their need. |
| 5: Accreditation | Laboratories should be United Kingdom Accreditation Services (UKAS) accredited and have evidence of External Quality Assessment (EQA), Internal Quality Control (IQC) and Internal Quality Assurance (IQA). |

According to click through data,[17] the majority of Google users do not go past the first page of search results, however for thoroughness the first five pages of results were screened by title and description. All Amazon results were screened. Inclusion criteria for a provider were:

1. The test had to be available in the UK
2. The test had to be either for self-sampling or self-testing
3. The test was not provided by an individual borough (administrative unit), as these are geographically limited and often use National Health System (NHS) services.

Data on the tests available, price, specimen type and advice/information were extracted iteratively from eligible websites. For products identified through third-party sellers, the original provider was identified and any other tests they provided also recorded.

### Provider questionnaire

Further information was requested from providers and associated laboratories through an online questionnaire, guided by categories identified during data extraction and BASHH guidelines.[7] Questionnaires were tailored for each provider, depending on the tests they provided and the information available on their website. The full set of questions is available in Supplementary Table 2. The questionnaire was first sent to providers in July 2020. In March 2021, BASHH published a position statement regarding online services. This highlighted inappropriate use of multiplex testing platforms, and suboptimal antibiotic treatment regimens for bacterial STIs,[12] emphasising the presence of inappropriate testing, incorrect specimens and poor treatment options that may have implications for AMR. The statement called for increased regulation of these services, as some providers’ practices are inconsistent with national guidelines. The questionnaire was sent again in April 2021 to the providers who did not respond in the first round, in the hope that the position statement publication would increase the response rate.

**Table 2:** Chart showing the pathogens tested for by online self-test and self-sample providers, and the samples requested for each pathogen. A full list of guidelines compared against is available in Supplementary Table 3.

| Type | Provider ID | Chlamydia | Gonorrhoea | Oral/Rectal chlamydia or gonorrhoea | Syphilis | HIV | Hepatitis B | Hepatitis C | Herpes (1&2) | Trichomoniasis | *Mycoplasma genitalium* | *Mycoplasma hominis* | *Ureaplasmas* | *Gardnerella* (as indicator of bacterial vaginosis) | *Haemophilus ducreyi* | Human Papillomavirus | Yeasts |
| --- | --- | --- | --- | --- | --- | --- | --- | --- | --- | --- | --- | --- | --- | --- | --- | --- | --- |
| BASHH Recommendation [7,16] |  | V (female), U (male), C if clinician collected | V (female), U (male) | S | B or lesion swab | B | B | B | Lesion swab or B | V, US, U | U (male and female), V/C (female) – symptomatic only | Not advised for routine testing | Not advised for routine testing | Not recommended as an indicator of bacterial vaginosis | Lesion swab – symptomatic only | Lesion Swab – symptomatic only (screening uses C which are clinician collected) | V – symptomatic only |
| Self-Test | 1 |  |  |  |  | B |  |  |  |  |  |  |  |  |  |  |  |
|  | 2 | U/C/US | U/C/US |  | B |  |  |  |  | C/US |  |  |  |  |  |  |  |
|  | 3 | C/US | C/US |  | B | B | B | B |  |  |  |  |  |  |  |  |  |
|  | 4 | C |  |  |  |  |  |  |  |  |  |  |  |  |  |  |  |
|  | 5 | S | S |  |  |  |  |  | B | V |  |  |  |  |  |  |  |
|  | 6 | U/C/US | ? |  | B | B/O | B | B |  |  |  |  |  |  |  |  |  |
|  | 7 |  |  |  |  |  |  |  |  | V |  |  |  |  |  |  |  |
|  | 8 | ? | S |  | B |  |  |  | B |  |  |  |  |  |  |  |  |
|  | 9 | V |  |  |  |  |  |  |  |  |  |  |  |  |  |  |  |
|  | 10 | C |  |  |  |  |  |  |  |  |  |  |  |  |  |  |  |
|  | 11 |  |  |  |  | B |  |  |  |  |  |  |  |  |  |  |  |
|  | 12 |  |  |  |  | B |  |  |  |  |  |  |  |  |  |  |  |
|  | 13 |  |  |  |  |  |  |  |  | ? |  |  |  | ? |  |  | ? |
| Self-Sample | 14 | U/V | U/V | S | B | B | B | B | U/V | U/V | U/V (Sp?) |  | U/V | U/V |  | V |  |
|  | 15 | U/V | U/V | S | B | B | B | B | U/V | U/V | U/V (Sp?) |  | U/V | U/V |  | V |  |
|  | 16 | U/V | U/V | S | B | B |  |  |  | U/V |  |  |  |  |  |  |  |
|  | 17† | U/V | U/V | S | B | B |  |  |  |  |  |  |  |  |  |  |  |
|  | 18 | U/V | U/V | S | B | B | B | B |  |  |  |  |  |  |  |  |  |
|  | 19† | U/V | U/V | S | B | B | B | B |  |  |  |  |  |  |  |  |  |
|  | 20 | U* | U* | S | B | B | ? | B | B/S/U | U | U (Sp?) |  | U | U |  | V |  |
|  | 21 | U/V | U/V | S | B | B |  |  | ? | ? | ? | ? | ? |  | ? | V |  |
|  | 22 | U | U |  | ? | ? | B | B | B | U | ? |  | ? | ? |  | V |  |
|  | 23 | U/V | U/V | S | B/S | B | B | B | B/S/U | U/V | U/V |  | U/V | U/V |  | V | V |
|  | 24 | U | U | S | B | B | B | B | B/U | U | U (Sp?) |  | U | U |  |  |  |
|  | 25† | U/V | U/V | S | B | B |  |  |  |  |  |  |  |  |  |  |  |
|  | 26† | U/V | U/V | S | B | B |  |  |  |  |  |  |  |  |  |  |  |
|  | 27 | U/V | U/V |  | B | B | B | B | U/V | U/V | U/V (Sp?) |  | U/V | U/V |  |  |  |
|  | 28 | U | U |  | B | B | B | B | B | ? | ? |  | ? | ? |  | V |  |
|  | 29 | U | U |  |  |  |  |  |  |  |  |  |  |  |  |  |  |
|  | 30 | U/V | U/V | S | B | B | B | B | U/V | U/V | U/V |  | U/V | U/V |  |  |  |
|  | 31† | U/V | U/V | S | B | B | B | B |  |  |  |  |  |  |  |  |  |

B = Blood; O=Oral transudate; S = Swab; Sp? = species unclear; U = Urine; US = urethral swab; V = Vaginal swab; ? = specimen unclear. This reflects where a provider explicitly states what sample is used. This is often not included on package tests, so may not reflect all tests offered; \* = most tests requested urine, but a vaginal swab was available separately. † Indicates a free service

### Comparison with national and regulatory diagnostic guidelines

Data obtained from provider services were categorised into: test type, specimen type, diagnostic accuracy, health information, follow-up/treatment, and accreditation. Comparison of services offered with BASHH guidelines was then conducted.[7,16] A full list of guidelines used for specific pathogens is available in Supplementary Table 3.

## RESULTS

### Overview of provider responses

The Google and Amazon search returned 13 self-test and 18 self-sample providers, as well as two laboratories that serviced multiple providers. In the first round of surveys, two providers completed the questionnaire, and one requested a phone call. The second round of surveys prompted four more replies. Therefore, most information was collected from provider websites. Provider names have been anonymised, in accordance with the survey terms of consent (Supplementary Table 2). The main guidelines providers were compared to are summarised in Table 1. Tests available and specimen type requested are shown in Table 2, compared to national guidelines. Further details of tests identified can be found in Supplementary Tables 3a and 3b. In general, the providers that were closest to the guidelines were free services, providing an appropriate range of tests, correct sample types, and up-to-date, comprehensive information.

### Test audience

Standard 1 was often not met. BASHH advice is that symptomatic users are not suitable for online services as they need to be examined,[7] yet providers specifically targeted this group and recommended a large panel of tests. Additionally, five self-test providers offered tests that were marked as being for professional use only. With regards to sexual history taking pre-testing, some providers used an online questionnaire to recommend the most appropriate options, but most providers did not have this feature.

### Test Process

Whilst providers did offer tests for the minimum requirement in standard 2, these were often available individually and in various packages, leaving users able to pick and choose. For the 13 self-testing kits, the main pathogens were chlamydia (n=8), HIV (n=5), and gonorrhoea (n=5). Less common pathogens were herpes (n=2), trichomoniasis (n=4), syphilis (n=4), hepatitis B (n=2), hepatitis C (n=2), *Gardnerella* (n=1) and *Candida albicans* (n=1). Currently, only HIV self-tests are approved for use in the UK,[18] which implies the availability of other self-tests is not regulated.

All 18 self-sample providers offered tests for chlamydia and gonorrhoea, but availability varied for other tests (Table 1). Self-sample tests were available in various combinations, with as many as 12 tests in one bundle. Free services provided a smaller range of tests than paid services. Ten self-sample services offered tests individually or within bundles for organisms generally regarded as commensal, such as *Ureaplasmas* or *Mycoplasmas*, however, it is recommended that these should not be routinely tested for.[14,15] It was also often unclear precisely which species was being referred to. *Gardnerella* infection was often used as a proxy for bacterial vaginosis, contrary to recommendations.[19] Private providers often exaggerated the pathogenicity and importance of testing for these commensal organisms. For example, two paid services claimed an advantage over the NHS by testing for organisms not included in routine testing, whilst exaggerating the long-term effects of the infections.[14]

Specimen type often conflicted with guidelines (Table 2). For self-test services, specimen type was appropriate where it could be identified, however five providers requested cervical samples, which should be clinician-collected.[20] As the test package inserts were unavailable, it was not possible to assess whether instructions on use were clear. For self-sample services, five providers only requested urine samples for chlamydia and gonorrhoea, with four of these having no option for females to order a vaginal swab, which is recommended.[20,21] The remaining provider did offer a vaginal swab, however this was stand-alone and offered separately from their main test package. Additionally, for herpes infections, at least seven self-sample providers requested urine samples. BASHH guidance states “Urine tests are inappropriate for the diagnosis of herpes” and instead recommends that lesion swabs are taken, or blood for serology.[22]

Eleven self-test providers reported sensitivity and specificity estimates, all of which were >85%, however information about reference tests used or sample sizes was often unavailable. Due to lack of website information and low survey response rate, it was not possible to obtain information on diagnostic accuracy from most self-sample providers. Those who reported these results (n=5) gave values >95% for sensitivity or specificity, but in three of these, only the HIV test performance was stated.

### Health information and signposting

To assess standard 3, we extracted information on whether sites gave information on STI symptoms, window periods, transmission details and health promotion (e.g. condom use). Health information was often lacking or inconsistent. For self-tests, as the package insert was only available for four providers, it was difficult to ascertain health promotion materials that may be provided to those purchasing the tests. As self-tests were often marked as professional use only, there could be lack of appropriate health information or signposting, as information would be targeted at a clinician. Using information from provider websites, only five self-sample providers provided information on window periods, transmission details and infection prevention. For other providers, information was not on the same page as the test or was inconsistently mentioned across different tests. One provider gave links to Wikipedia. Advice on accessing HIV post-exposure prophylaxis (PEP) was also not mentioned by eight of the self-sample providers; guidelines stipulate that users in need of this service should be directed to a clinic.[7]

### Follow-up/treatment

It was difficult to assess whether standard 4 was met as often post-diagnosis processes were not shared. Where post-positive diagnosis information was provided, options involved a private consultation, treatment ordered online (mainly for chlamydia), or advice to visit a General Practitioner (GP). Partner notification was often mentioned non-specifically and may instead have been discussed post-diagnosis. Exact treatment options were unclear, however at least one provider offered an oral course of azithromycin and cefixime for gonorrhoea treatment which was easy to purchase online, instead of the recommended intramuscular ceftriaxone as per BASHH recommendations, which requires a visit to a healthcare professional.[21]

### Accreditation

Although the BASHH standards do not refer to accreditation for self-tests, it is recommended that they hold the CE-mark.[18] Eleven self-test providers had at least one of their tests CE-marked, two claimed World Health Organization (WHO) approval and one claimed Food and Drug Administration (FDA) accreditation. One self-test provider marked their chlamydia and gonorrhoea self-tests with an NHS logo, describing themselves as an NHS provider, but whether that product had received NHS endorsement was unclear. For self-sample providers, United Kingdom Accreditation Service (UKAS) accreditation was claimed as required in Standard 5, however, it was often used as a blanket term for the laboratory without details of the specific laboratory service that had received accreditation.[23] Of the two laboratory providers we were able to identify, one had UKAS accreditation for certain STI tests and the other had UKAS accreditation for tests other than STIs. Care Quality Commission (CQC) accreditation was reported for 12 self-sample providers, although mostly only for laboratories used for sample testing, as opposed to the providers themselves.

## DISCUSSION

This study identified and analysed 31 providers of online tests in the UK. We found significant areas of suboptimal service that often conflicted with national guidelines on STI diagnostics and management. These included a lack of health promotion information, lack of sexual history taking, use of tests licensed for professional use only marketed for self-testing, inappropriate infections tested for, incorrect specimen type used, and lack of advice for post-diagnosis management. As a result, users are at risk of taking unnecessary tests, with poor performance, that could lead to incorrect results, inappropriate management and receiving inadequate clinical information and support.

The first limitation of this study was low questionnaire response rate from providers, despite a follow-up in 2021 following the BASHH position statement publication,[12] meaning that not all aspects of care could be evaluated. Data considered missing in our analysis may have been available once the user had bought the test. Furthermore, data were extracted from websites in July 2020 but providers may have subsequently updated their websites. Although our internet search was comprehensive, it is not possible to identify all online STI test providers. The sample analysed here may therefore not be fully representative of all providers. This lack of representativeness may be further compounded by the small number of providers who responded to the survey and for whom we therefore have more extensive data. However, the low response rate we observed has been seen in similar studies where providers were contacted for information, and is not unique to our study.[24,25] In addition, as our study was unfunded, tests could not be purchased to identify whether information was available post-purchase. This also meant we were unable to test the services independently, either from a user perspective through a “mystery shopper” exercise,[26] or from a diagnostic accuracy perspective by independently assessing test performance claims. These are obvious next steps for future work. Despite this, we were able to collect large amounts of information from provider websites, giving an accurate perspective of what a consumer would experience when choosing to order an online test.

Whilst it was difficult to assess test performance in the identified providers due to lack of available information and inability to perform independent evaluations, it is expected that test performance was sub-optimal in at least some instances.[27] BASHH guidelines note that chlamydia and gonorrhoea tests should be nucleic acid amplification test (NAAT)-based. [20,21] However, many non-NAAT chlamydia and gonorrhoea self-tests were on offer. Although many products had CE-marks, as noted by BASHH,[12] this is easily obtainable and tests may not have been adequately validated. Using incorrect sample types, or being sold tests approved for professional use only, as seen in our evaluation, may exacerbate poor test performance and add to this issue.[20–22]

The lack of appropriate health information given by providers poses a risk to users on multiple levels. Access to healthcare professionals as part of online STI services is recognised as important for offering information, technical assistance, and support such as reassurance and navigating relationship implications.[28] Receiving accurate information regarding appropriate services and tests is critical to providing appropriate patient care, ensuring that patients receive the correct tests relevant to their situation. In contradiction to this, we found that several online providers specifically targeted symptomatic patients, whereas standard BASHH guidelines are clear that online services are appropriate for asymptomatic individuals only,[7] as well as not being signposted to vital services such as PEP for HIV.[7] Patients were also frequently offered testing for commensal *Mycoplasmas* and *Ureaplasmas*,[14] which could lead to unnecessary costs, treatments, and results of uncertain significance,[15,29] resulting in emotional distress and poor antimicrobial stewardship. These additional tests were only found in paying providers’ services, suggesting that they may be more motivated by profit than by high quality healthcare provision.[30] It would be important to understand why individuals choose to pay for testing rather than opting for the free services, to help ensure patients are offered the best possible care.

Whilst this is the first time to our knowledge that an evaluation of UK online testing providers has been conducted, studies in other countries externally assessing online test providers have reported similar results. A 2010 study of online tests in America was able to perform independent assessments of online STI test providers, finding that they were hard to contact, and although self-tests had poor performance, self-sample tests had high accuracy.[27] Providers of chlamydia online tests in The Netherlands were found to often not meet quality indicators regarding health promotion or follow-up (especially for self-tests), but the process of an evaluation taking place did provoke providers to improve their service.[25] Similarly, an Australian study of HIV self-tests showed that none conformed to national product guidelines, and often had inadequate pre-test information and linkage to care.[24] These studies demonstrate that sub-optimal online testing service provision is a problem across the world. Actions such as publications highlighting short-falls and position statements with recommendations may create short-term impacts. However, if there are no mechanisms to maintain improved practice and prevent providers from, for example, appearing under a different name,[31] these efforts are of little long-term benefit. For there to be sustained improvements in patient care, regulatory change is needed so that providers are regularly monitored and can be held to account. Part of this involves services frequently being evaluated against national guidelines, which must also be continually updated to adapt to evolving service provision.[8,12]

## CONCLUSION

Online testing is a welcome addition to STI diagnostics, offering a convenient and flexible option for users. However, the proliferation of providers that do not follow guidelines, in particular for-profit sites, jeopardises these advantages and puts users at risk. If current trends continue, online testing usage will increase, resulting in more online providers as demand increases. Regulatory change is required to ensure that the standard of care received online meets national guidelines to protect patients and the wider population from the repercussions of underperforming or inappropriate tests. If we do not act now, patients will continue to receive sub-optimal care with potentially significant adverse personal, clinical, and public health implications.

## Data Availability

The data that support the findings of this study are available upon reasonable request from the corresponding author, [EMHE]. Some data are not publicly available because participants of this study did not agree for their data to be shared publicly

## Acknowledgments

We thank all providers who responded to the questionnaire. We would also like to thank the BASHH Bacterial Special Interest Group (BSIG) members for reviewing the manuscript.

EMHE would like to thank Julius Schachter who set the challenge to address this issue at the 8^th^ Meeting of the European Society for Chlamydia Research, September 2016, Oxford, UK. It is not yet the outcome you hoped for, but progress is slowly being made.

## Funding and all other required statements

KMET acknowledges support from HDRUK CFC 0129 and PH and KMET acknowledge support from the Health Protection Research Unit in Behavioural Science and Evaluation, NIHR 200877, at University of Bristol.

