## Supplementary Data for "Online tests for sexually transmitted infections – Friend or Foe? An analysis of providers in the United Kingdom"

Supplementary Table 1

***Supplementary Table 1:*** *Search strategy used in Google. SS = self-sample provider, ST = self-test provider. For each search (1-5) the search terms for each concept were combined with the Boolean operator AND. When the acronyms STI (sexually transmitted infection) and STD (sexually transmitted disease) were included in their full form, results were mainly educational or medical, so they were not included in the search. The results for each were listed and pooled to produce a final list of providers. Amazon search terms were similar, but searches were run individually without Boolean operators.*

| Search Number | Search Concepts (combined with AND) | | | Number of Providers Identified |
| --- | --- | --- | --- | --- |
|  | Online | STI | Test |  |
| 1 | home OR online OR instant OR rapid | sti OR std | test | 20  (15 SS, 5 ST) |
| 2 | home OR online OR instant OR rapid | chlamydia OR gonorrhoea OR syphilis OR hiv OR herpes | test | 16 (11 SS, 5 ST) |
| 3 | home OR online OR instant OR rapid | sti OR std | test OR diagnosis OR diagnostic | 19 (15 SS, 4 ST) |
| 4 | home OR online OR instant OR rapid | chlamydia OR gonorrhoea OR syphilis OR hiv OR herpes | test OR diagnosis OR diagnostic | 16 (11 SS, 5ST) |
| 5 | home OR online OR instant OR rapid | sti OR std OR chlamydia OR gonorrhoea OR syphilis OR hiv OR herpes | test OR diagnosis OR diagnostic | 13  (11 SS, 2 ST) |

Supplementary data 2 – Questionnaires

Questionnaires were adapted to each provider, depending on the tests they provide and the information already available. For questions pertaining to specific test details, and where providers had more than one test, all tests were listed where the template specifies [Test Name/Pathogen]. All questionnaires included the following consent statement:

*Dear Sir/Madam,*

*Thank you for taking part. The information collected during this survey will be used in a research project investigating the quality and availability of online tests for sexually transmitted infections (STIs), taking place at the London School of Hygiene & Tropical Medicine.*

*This survey is in reference to your diagnostic test [test description and website link]. By providing information, you are consenting to have that information included in this project. No identifying information such as the company name will be included in any published work, and any information collected will be securely stored by the study team. If you wish to withdraw consent at any time after completing the survey, please e-mail [e-mail address] (study lead).*

*The survey is separated into three parts, 1) pre-test information 2) test performance and 3) post-test information. All questions are optional.*

*You may also reply in email or document form to [e-mail address] if you would prefer, and do not hesitate to email any questions about the survey or the project.*

**2a: Self-Test Provider Questionnaire**

|  | Question Text | Answer Options | | | | | | | | | |
| --- | --- | --- | --- | --- | --- | --- | --- | --- | --- | --- | --- |
| 1 | Is the [test details] suitable for someone who has symptoms of an STI? | 🞎Yes | | | | | | | | | |
|  |  | 🞎No | | | | | | | | | |
|  |  | 🞎Other (Please Specify) | | | | | | | | | |
| 2 | Is the [test details] suitable for home use by someone who is not a clinical professional? | 🞎Yes | | | | | | | | | |
|  |  | 🞎No | | | | | | | | | |
| 3 | What window period (time between being infected and the infection being detectable by the test) do you advise before using the [test detail] |  | Window Period (days) | | | | | | | | |
|  |  | [Test Name/Pathogen] | *Free text* | | | | | | | | |
| 4 | What accreditation does your test have (e.g. CE mark, ISO)? | *Free text* | | | | | | | | | |
| 5 | What specimen does your test require? (Tick all that apply) |  | Urine (male) | Urine (female) | | Vaginal Swab | Cervical Swab | Blood | Urethral Swab | | Other |
|  |  | [Test Name/Pathogen] | 🞎 | 🞎 | | 🞎 | 🞎 | 🞎 | 🞎 | | Free text |
| 6 | Please describe the target and mechanism used for your diagnostic test |  | What is the diagnostic target for this pathogen in your test? e.g. antigen, antibody | | | | What is the mechanism of this test? (e.g. lateral flow immunochromatographic assay) | | | | |
|  |  | [Test Name/Pathogen] | *Free text* | | | | *Free text* | | | | |
| 7 | Please provide information on the specificity and sensitivity of your test and how it was calculated. For reference: Sensitivity = true positives/true positives + false negatives Specificity = true negatives/true negatives + false positives If you do not have access to this information please discuss it with someone who does, or provide their details and we can contact them directly. If it is easier to provide this in document form, please email it to [student email address] |  | Sample Type | | Sensitivity (%) | Sensitivity Calculation | Specificity (%) | Specificity Calculation | | Reference Test | |
|  |  | [Test Name/Pathogen] | *Free text* | | *Free text* | *Free text* | *Free text* | *Free text* | | *Free text* | |
| 8 | Please provide details of any publications or other literature describing the test's use and/or performance: | *Free text* | | | | | | | | | |
| 9 | What advice is given to the user after a negative test result? | *Free text* | | | | | | | | | |
| 10 | If a test is positive, what advice is given to the user for the next stage of care? (e.g. confirmatory tests, treatment options, follow up with medical professional) | *Free text* | | | | | | | | | |
| 11 | If a diagnosis is positive, what advice is given in regard to partner notification (including look back period)? | *Free text* | | | | | | | | | |
| 12 | Is any other advice is given after a positive test result? | *Free text* | | | | | | | | | |
| 13 | Is there any other information you would like to include? | *Free text* | | | | | | | | | |

**2b: Self-Sample Provider Questionnaire**

|  | Question Text | Answer Options | | | | | | | | | | | | | | | |
| --- | --- | --- | --- | --- | --- | --- | --- | --- | --- | --- | --- | --- | --- | --- | --- | --- | --- |
| 1 | Are any checks of eligibility done before providing a test to ensure the test is appropriate to the user (e.g. a patient questionnaire relating to symptoms, risk behaviours or previous diagnoses)? | 🞎Yes | | | | | | | | | | | | | | | |
|  |  | 🞎No | | | | | | | | | | | | | | | |
|  |  | If yes, please provide details:  *Free text* | | | | | | | | | | | | | | | |
| 2 | Is your service suitable for users that have symptoms of an STI? | 🞎Yes | | | | | | | | | | | | | | | |
|  |  | 🞎No | | | | | | | | | | | | | | | |
| 3 | What national guidelines do you follow for running your service (e.g. BASHH)? | *Free text* | | | | | | | | | | | | | | | |
| 4 | What window period (time between being infected and the infection being detectable by the test) do you advise before using the [test detail] |  | | Window Period (days) | | | | | | | | | | | | | |
|  |  | [Test Name/Pathogen] | | *Free text* | | | | | | | | | | | | | |
| 5 | What laboratory do you use for your diagnostic tests and what accreditations do they have (e.g. UKAS, ISO)? | *Free text* | | | | | | | | | | | | | | | |
| 6 | Which of the following species of Mycoplasma and Ureaplasma do you test for? | *Mycoplasma hominis* | *Mycoplasma genitalium* | | | | *Ureaplasma parvum* | | | *Ureaplasma urealyticum* | | | *Ureaplasma (not species specific)* | | | *Other* | |
|  |  | 🞎 | 🞎 | | | | 🞎 | | | 🞎 | | | 🞎 | | | *Free text* | |
| 7 | Do any of your tests include antimicrobial susceptibility testing? | 🞎 Yes | | | | | | | | | | | | | | | |
|  |  | 🞎 No | | | | | | | | | | | | | | | |
|  |  | If yes, please provide details: *Free text* | | | | | | | | | | | | | | | |
| 8 | What samples do you use to test for the following pathogens in the [test package name]? Tick all that apply. |  | | Urine (male) | Urine (female) | | | Oral Swab | Rectal Swab | | Vaginal Swab | Cervical Swab | | Blood | Urethral Swab | | Other |
|  |  | [Test Name/Pathogen] | | 🞎 | 🞎 | | | 🞎 | 🞎 | | 🞎 | 🞎 | | 🞎 | 🞎 | | *Free text* |
| 9 | Please describe the target and platform/assay used for your diagnostic tests. If any tests are done in combination, or if there are multiple tests used, please include that in the details. If you do not have access to this information, please discuss it with someone who does or provide their details and we can contact them directly. For HIV, please also specify the generation of test if applicable. |  | | What is the diagnostic target for this pathogen? e.g. antibody, nucleic acid | | | | | | | | What platform or assay is used to run this test? | | | | | |
|  |  | [Test Name/Pathogen] | | *Free text* | | | | | | | | *Free text* | | | | | |
| 10 | Please provide information on the specificity and sensitivity of your test and how it was calculated. For reference: Sensitivity = true positives/true positives + false negatives Specificity = true negatives/true negatives + false positives If you do not have access to this information please discuss it with someone who does, or provide their details and we can contact them directly. If it is easier to provide this in document form, please email it to [student email address] |  | | Sample Type | | Sensitivity (%) | | | Sensitivity Calculation | | | Specificity (%) | | Specificity Calculation | | Reference Test | |
|  |  | [Test Name/Pathogen] | | *Free text* | | *Free text* | | | *Free text* | | | *Free text* | | *Free text* | | *Free text* | |
| 11 | Please provide details of any publications or other literature describing the test's use and/or performance: | *Free text* | | | | | | | | | | | | | | | |
| 12 | What advice is given to the user after a negative test result? | *Free text* | | | | | | | | | | | | | | | |
| 13 | What is the next stage of care when the user tests positive for the infections listed below (e.g. treatment is provided online, user is referred to a sexual health clinic, confirmatory testing is required), and what advice about partner notification is given (including look back period)? |  | | | | | Next stage of care | | | | | | Advice/recommendations about partner notification | | | | |
|  |  | [Test Name/Pathogen] | | | | | *Free text* | | | | | | *Free text* | | | | |
| 14 | Do you recommend any tests be repeated at a later date? | 🞎 Yes | | | | | | | | | | | | | | | |
|  |  | 🞎 No | | | | | | | | | | | | | | | |
|  |  | If yes, please provide details of what recommendations are made: *Free text* | | | | | | | | | | | | | | | |
| 15 | If a chlamydia test is positive, what is your advice to the user regarding testing for lymphogranuloma venereum (LGV)? | *Free text* | | | | | | | | | | | | | | | |
| 16 | Please describe any other advice given after a positive diagnosis from any of your tests: | *Free text* | | | | | | | | | | | | | | | |
| 17 | Do you report your results to any external body (e.g. Public Health England) for surveillance purposes? | 🞎 Yes | | | | | | | | | | | | | | | |
|  |  | 🞎 No | | | | | | | | | | | | | | | |
|  |  | If yes, please provide details: *Free text* | | | | | | | | | | | | | | | |
| 18 | Is there any other information you would like to include? | *Free text* | | | | | | | | | | | | | | | |

Supplementary Tables 3a and 3b

Providers were compared to pathogen specific guidelines where available, and other literature where guidelines have not been published: chlamydia[1], gonorrhoea,[2] syphilis,[3] HIV,[4] hepatitis,[5] herpes,[6] trichomoniasis,[7] Mycoplasma genitalium,[8] Mycoplasma hominis,[9,10] Ureaplasmas,[9,10] Gardnerella,[11] chancroid,[12] human papillomavirus,[13,14] yeasts.[15]

***Supplementary Table 3a:*** *Description of the characteristics of self-test kits for sexually transmitted infections found available online.*

| **Provider** | **Infection** | **Sample Type Requested** | **Positive Diagnosis Advice** | **Professional Use Only?** | **Accreditation** | **Sensitivity (sample type)** | **Specificity (sample type)** | **Reference Test** |
| --- | --- | --- | --- | --- | --- | --- | --- | --- |
| 1* | HIV | Blood | Consult a doctor for confirmatory testing | No | CE, WHO approved | 100% (Blood) | 99.8% (Blood) | PCR |
| 2* (answers reflect survey response, which differed to information available on the website) | Chlamydia | Cervical/urethral swab/Urine | Seek confirmatory testing and treatment for you and your partner | Yes | CE | 91.3% (Swab/Urine) | 98.1% (Swab/Urine) | PCR |
|  | Gonorrhoea | Cervical/urethral swab |  | Yes | CE | 97% (Swab) | 96% (Swab) | Culture |
|  | Trichomoniasis | Cervical/urethral swab |  | Yes | CE | 85.7% (Swab) | 97.5% (Swab) | Another rapid test (Unnamed) |
|  | Syphilis | Blood |  | Yes | CE | 99.7% (Blood) | >99.9% (Blood) | TPPA |
| 3 | Chlamydia | Cervical/urethral swab | / | Yes | CE | 90% (Cervical Swab) 80.9% (Male Urethral Swab) 92.3% (Male Urine) | 96.5% (Cervical Swab) 94.3% (Male Urethral Swab) >99.9% (Male Urine) | PCR |
|  | Syphilis | Blood |  | Yes | CE | >99.9% (Whole Blood) | 99.7% (Whole Blood) | / |
|  | Gonorrhoea | Cervical/urethral swab |  | Yes | CE | 90.9% (Cervical Swab) 90% (Male Urethral Swab) | 96.4% (Cervical Swab) 96.8% (Male Urethral Swab) | Culture |
|  | Hepatitis B Surface Antigen | Blood |  | Yes | / | >99.9% (Whole Blood) | 99.3% (Whole Blood) | / |
|  | Hepatitis C | Blood |  | / | / | 99.1% (Whole Blood) | 99.5% (Whole Blood) | / |
|  | HIV Antigen/antibody | Blood |  | Yes | / | >99.9% | 99.5% | / |
| 4 | Chlamydia | Cervical Swab (advertised as vaginal) | See a health professional | No | CE | 85.7% (Cervical Swab) | 98.3% (Cervical Swab) | PCR |
| 5 | Chlamydia | Swab (source unclear) | See a health professional | Yes | CE, FDA | 98.5% "accurate" | | / |
|  | Gonorrhoea | Swab (source unclear) |  | Yes | CE, FDA | 98.5% "accurate" | | / |
|  | Genital Herpes (HSV2) | Blood |  | Yes | CE, FDA | 99% "accurate" | | / |
|  | Oral Herpes (HSV1) | Blood |  | Yes | CE, FDA | 99% "accurate" | | / |
|  | Trichomoniasis | Vaginal swab |  | Yes | CE, FDA | 98.5% "accurate" | | / |
| 6 | Chlamydia | Cervical/urethral swab/urine | / | Yes | CE | / | / | / |
|  | Gonorrhoea | / |  | Yes | CE | / | / | / |
|  | Syphilis | Blood |  | Yes | CE | / | / | / |
|  | HIV (Blood) | Blood |  | Yes | WHO Prequalified | / | / | / |
|  | HIV (Oral) | Oral transudate |  | Yes | / | / | / | / |
|  | Hepatitis B Surface Antibody | Blood |  | Yes | / | 97.30% | 99.20% | / |
|  | Hepatitis B Surface Antigen | Blood |  | Yes | / | / | / | / |
|  | Hepatitis C | Blood |  | Yes | / | 99% | 99.80% | / |
| 7* | Trichomoniasis | Vaginal swab | See a health professional | No | CE | 99% | 100% | Culture |
| 8 | Chlamydia | / | / | No | CE | / | / | / |
|  | Syphilis | Blood |  | No | / | / | / | / |
|  | Gonorrhoea | Swab (source unclear) |  | Yes | CE | / | / | / |
|  | HSV1 | Blood |  | Yes | CE | / | / | / |
|  | HSV2 | Blood |  | Yes | CE | / | / | / |
| 9 | Chlamydia | Vaginal swab | / | No | / | 98.3% | / | / |
| 10* | Chlamydia | Cervical Swab (advertised as vaginal) | See a health professional | No | CE | 85.7 % (Cervical Swab) | 98.3 % (Cervical Swab) | PCR |
| 11 | HIV | Blood | See a health professional for confirmatory tests | No | CE | 99.6% (Blood) | / | / |
| 12 | HIV | Blood | See a health professional for confirmatory tests | No | CE | 99.7% (Blood) | 99.9% (Blood) | Enzyme immunoassay and western blot |
| 13 | Gardnerella | / | / | No | / | 98.5% | 98.6% | PCR |
|  | Trichomoniasis | / |  | No | / | 100% | 99% | Wet mount microscopy and culture |
|  | Candida Albicans | / |  | No | / | 95.5% | 98.4% | Wet mount microscopy and culture |

*/ Indicates the information was not stated or unclear. HSV = herpes simplex virus, TPPA = Treponema pallidum particle agglutination assay, FDA = food and drug administration, WHO = World Health Organisation. * Indicates the provider responded to the survey.*

***Supplementary Table 3b:*** *Description of the characteristics of self-sample provider websites and services.*

| **Provider** | **Window Period Stated** | **Advised for symptomatic users?** | **Pre-test Screen** | **Symptom Information** | **Transmission Information** | **Health Protection Information** | **HIV PEP signpost** | **Positive Result Guidance** | **Accreditations** | **Price Range** |
| --- | --- | --- | --- | --- | --- | --- | --- | --- | --- | --- |
| 14 | Yes | Unclear | No | Yes | Inconsistently | Condoms mentioned on two test pages, not all | Yes | Treat online or arrange a consultation | CQC | £29-£244 |
| 15 | Yes | Unclear | No | Yes | Inconsistently | Thorough description on one test page but not all | Yes | Treat online or arrange a consultation | CQC | £27.99-£225.99 |
| 16 | Yes | No | Yes | Yes, not on the test page | Yes, not on the test page | Yes, not on the test page | Yes | Treat online or referral | Unclear | £27.99-£99.95 |
| 17* | Yes | No | Yes | Yes | Yes | Yes | Yes | Refer to treatment | CQC, Claims a UKAS Accredited Laboratory | Free |
| 18 | Yes | No | Yes | Yes | Yes | Yes | Yes | Treat online/referral | Unclear | £28-£128 |
| 19 | Yes | No | Yes | Yes | Yes | Yes | Yes | Treat online/referral | CQC, UKAS accredited laboratory | Free |
| 20 | Yes | Unclear | No | Yes, not on the test page | Yes, not on the test page | Condoms mentioned on one test page, not all | No | Treat online, consultation or referral | Unclear – UKAS badge given for quality management service, not tests provided | £35-£299 |
| 21 | No | No | No | Yes, not for all pathogens | Yes, not for all pathogens | Links to Wikipedia | No | Signpost to local services | CQC, UKAS accredited laboratory but not accredited for all pathogens tested for | £19.99-£114.99 |
| 22 | Yes | Yes | No | No | No | No | No | Results will be discussed with the user | Unclear | £49-£209 |
| 23 | Yes, not for all pathogens | Yes | No | Yes, not on the test page | Yes, not on the test page and not for all pathogens | No | No | Treat online or a consultation or referral | CQC | £29.95-£299.95 |
| 24 | Yes, not for all pathogens | Unclear | Limited | Yes, not for all pathogens and not always on the test page | Yes, not for all pathogens and not always on the test page | Condoms and safe toy use mentioned on each STI info page | No | Phone consultation | Claim they use a UKAS accredited laboratory but no further details to verify this | £95-£239 |
| 25* | Yes | Yes (survey), No (website) | Yes | Yes | Yes | Yes | Yes | Consultation | Unclear | Free |
| 26 | Yes, not for all pathogens | Unclear | No | Yes | Yes | No | No | Unclear | CQC, UKAS accredited laboratory | Free |
| 27 | No | Unclear | No | Yes, not for all pathogens | Yes, not for all pathogens | No | No | Consultation | CQC, | £34-£225 |
| 28 | Yes, not for all pathogens | Yes | No | Yes, not for all pathogens | Yes, not for all pathogens | No | No | Advised to see your doctor, states they will not diagnose or consult | CQC, claims UKAS laboratory however this lab is not accredited for STIs. May be accredited for other services they provide | £37-£251 |
| 29 | Yes | Unclear | No | Yes, not for all pathogens | No | Condoms advised | N/A (no HIV test) | Treat online/referral | CQC | £35 |
| 30 | Yes | Unclear | No | Inconsistently | Yes, not for all pathogens | Condoms and safe toy usage mentioned on some pages but not all | Yes | Treat online/referral | CQC | £32-£200 |
| 31* | Yes | No | Yes | Yes | Yes | Yes | Yes | Treat online/referral | CQC, UKAS Accredited Laboratory | Free |

*CQC = care quality commission, PEP = post-exposure prophylaxis, ISO = international organisation for standardization, UKAS = United Kingdom Accreditation Service. * indicates that the provider responded to the survey*
